# Association study of *BDNF* Val66Met gene polymorphism with bipolar disorder and lithium treatment response in Indian population

**DOI:** 10.1101/2020.09.28.20203513

**Authors:** Pradip Paul, Ravi Kumar Nadella, Somdatta Sen, Dhruva Ithal, Jayant Mahadevan, Y C Janardhan Reddy, Sanjeev Jain, Meera Purushottam, Biju Viswanath

## Abstract

**Background:** The association of the Val66Met (rs6265) polymorphism in the brain derived neurotrophic factor (*BDNF*) gene with bipolar disorder (BD) and response to lithium treatment has been suggested, though inconsistently. The considerable diversity of allele frequency across different populations contributes to this. There is no data from South Asia till date. Hence, we examined the association of this polymorphism in BD cases from India, and its correlation with lithium treatment response.

**Methods:** BD patients (N=301) were recruited from the clinical services of National Institute of Mental Health and Neurosciences (NIMHANS), India. Lithium treatment response for 190 BD subjects was assessed using Alda scale by NIMH life charts. Patients with total score ≥ 7 were defined as lithium responders (N=115) and patients with score <7 were defined as lithium non-responders (N=75). Healthy controls (N = 484) with no lifetime history of neuropsychiatric illness, or a family history of mental illness were recruited as control set. Genotyping was performed by TaqMan genotyping assay.

**Results:** Genotype and allele frequency of *BDNF* Val66Met SNP was significantly different (χ2 = 7.78, p=0.02) in cases compared to controls, and the Val(G) allele was more frequent (χ2 = 7.08, p=0.008) in BD patients. Although there was trend towards significance for greater occurrence of Val(G) allele among lithium non-responders, this difference did not persist in an analysis of extreme response groups.

**Conclusions:** The Val(G) allele of *BDNF* Val66Met polymorphism is associated with risk of BD in this sample, but is not related to response to lithium.

## Introduction

Bipolar disorder (BD), which affects 1-4% of the world’s population (Scaini et al., 2020), is a severe and chronic psychiatric illness characterized by episodes of depression alternating with mania or hypomania. With a mean age of onset of 18-20 years (Merikangas et al., 2011), BD is often associated with cognitive and functional impairment which pose a great burden on patients and families (Grande et al., 2016). There is also raised mortality with approximately 20% of patients committing suicide over the course of illness (Berkol et al., 2016).

Family and twin studies have shown that genetic factors play a significant role in the development of the illness across various populations studied (Escamilla and Zavala, 2008), with concordance rate of 38.5 - 43% for monozygotic twins and 4.5 – 5.6% for dizygotic twins (Gordovez and McMahon, 2020). The heritability of BD is estimated to be between 60% and 80% (Kerner, 2014).

The brain derived neurotrophic factor (*BDNF*) gene has been one of the well-studied genes that have been shown to be associated with BD (Mandolini et al., 2019). The *BDNF* gene maps on chromosome 11p13 (Tsai, 2018). The nonsynonymous G/A SNP (rs6265) at the position 196 of exon 2 results in substitution of valine (Val) to methionine (Met) (Val66Met) (Shen et al., 2018); and the variation at this locus interacts with the action of mood-stabilizing treatments. Met allele attenuates the activity□dependent intracellular trafficking and secretion of neuronal BDNF (Björkholm and Monteggia, 2016; Machado-Vieira, 2018). A number of genetic studies have thus examined the effect of this *BDNF* Val66Met polymorphism on brain function and behavior, and neuropsychiatric illnesses including BD (Mandolini et al., 2019; Park et al., 2017; Rakofsky et al., 2012).

In a family based association study, the Val allele was significantly more transmitted in BD (De Luca et al., 2008). A genetic database for BD, documented 38 candidate gene association studies (2002-2015) related to BD and the *BDNF* Val66Met in different populations. Out of the 38 reports, 18 reported positive association of either variant (Val or Met) with BD (Chang et al., 2013). Val allele was suggested to be associated in Caucasian populations (Frazier et al., 2014) whereas findings in Han Chinese population showed Met allele to be associated with BD (Wang et al., 2012; Xu et al., 2010). While some meta-analyses on the European population have shown the Val allele to be associated with BD, there have been mixed results in other populations (González-Castro et al., 2015; Li et al., 2016; Rai et al., 2019).

The mood stabilizer lithium is known to modulate many biochemical pathways, such as neurotrophic signaling, where BDNF plays a prominent role (Machado-Vieira, 2018). Pharmacogenetic association experiments of lithium prophylaxis have shown that the Val allele is associated with poor response to lithium (Dmitrzak-Weglarz et al., 2008; Rybakowski et al., 2007, 2012). Wang et. al. (2012) reported that this allele is associated with poorer response to lithium in BD type 1, while in type 2 BD it is associated with good response (Wang et al., 2012). Other studies, however, have not been able to confirm these results (Masui et al., 2006; Michelon et al., 2006; Wang et al., 2014).

Overall, these findings indicate that the association of this polymorphism with BD risk, and lithium treatment response, may vary across populations. This ambiguity might also be due to differences in the frequency of the *BDNF*-Met allele across populations, as it varies from 0 to 72% (Tsai, 2018). There are no studies from South Asia till date. It may therefore be useful to evaluate the effect of this polymorphism in BD cases from India, and its correlation with lithium response.

## Methodology

### Sample selection

The study participants were 301 BD patients, who provided written informed consent, from the clinical psychiatry services of National Institute of Mental Health and Neurosciences (NIMHANS), India. The Institutional Ethics Committee (IEC), NIMHANS had approved the study protocol (NIMHANS IEC No.-Sl. No. 11, Behavioral Science; dated 12^th^ Aug, 2013). The diagnosis of BD was confirmed by 2 qualified psychiatrists independently according to DSM-IV-TR (American Psychiatric Association, 2000). For comparison, we identified 484 healthy controls (HC) also from south India who had no lifetime history of neuropsychiatric illness or a family history of mental illness in two generations.

Treatment response was ascertained as described in our earlier publication (Kapur et al., 2018), and was available for 190 subjects. This included assessments on the Alda Scale (Manchia et al., 2013) and NIMH life charts (Roy-Byrne et al., 1985). Lithium response was defined as a dichotomous phenotype, patients with score ≥ 7 were defined as lithium responders and patients with score <7 were defined as lithium non-responders (Hou et al., 2016; Kapur et al., 2018).

### Genotyping

Genomic DNA was extracted from peripheral blood leukocytes by sodium chloride precipitation (Miller et al., 1988) and quality and quantity of DNA was assessed by NanoDrop ND-100 (Thermo Scientific) spectrophotometer. Genotyping at rs6265 was performed by TaqMan genotyping assay (#C_11592758) from Thermo Fisher Scientific using Applied Biosystem 7500 Real time PCR Systems. The results were analyzed based on the amplification plot by using Applied Biosystem 7500 software version 2.0.5. Genotyping of 5% samples were confirmed by Sanger sequencing method using BigDye Terminator v3.1 Cycle Sequencing Kit (Thermo Fisher Scientific). The sequencing reaction were analyzed in 3500xL Genetic Analyzer, capillary electrophoresis using Sequencing Analysis Software (Thermo Fisher Scientific).

### Statistics

Chi-square test and Mann-Whitney tests were applied to assess the basic clinical characteristics. Chi-square / Fisher exact tests were carried out to explore the association of the polymorphism with BD and lithium treatment response. The tests were performed using GraphPad Prism version 7.00 for Windows, GraphPad Software, La Jolla California, USA, www.graphpad.com. The Hardy-Weinberg equilibrium was tested by comparing observed and expected genotype frequencies by two tailed chi square statistics using web-tool (Rodriguez et al., 2009). The statistical significance was set at p ≤ 0.05.

## Results

The basic clinical characteristics of controls, lithium responders and lithium non-responders has been described in Table 1. The *BDNF* Val66Met genotype and allele frequency results are tabulated and presented in Table 2. The genotype frequencies were in Hardy–Weinberg equilibrium. The genotype and allele frequency of Val66Met SNP was significantly different (χ^2^ = 7.78, df = 2, p=0.02) in cases compared to controls. The Val(G) allele was significantly higher (χ^2^ = 7.08, df = 1, p=0.008) in BD patients compared to controls with odds ratio (OR) = 1.47 and confidence interval (CI) = 1.11 to 1.96. There was no significant difference in genotype frequency of the Val66Met polymorphism between the lithium responders and non-responders (χ^2^ = 4.56, df = 2, p=0.56). We observed a non-significant trend towards an increased occurrence of the Val(G) allele among lithium non-responders in this analysis (χ2 = 2.71, df = 1, p=0.09). However, additional Chi-square analysis comparing BD subjects at the extreme ends of the distribution (total score ≥7 as lithium responder, N=115; and score ≤3 as non-responder, N=35) was performed, and this trend was not substantiated (χ2 = 2.1, df = 1, p=0.15).

**Table 1:** Basic clinical characteristics of study particpants:

| **Clinical variables** | **Cases**<br>**(N=301)** | **Controls**<br>**(N=484)** | **p-value** | **Lithium**<br>**responders**<br>**(N= 115)** | **Lithium non-**<br>**responders**<br>**(N=75)** | **p-value** |
| --- | --- | --- | --- | --- | --- | --- |
| **Age at assessment**<br>**(years) (Mean ± SD)** | 33.8 ± 11.4 | 39.1 ± 18 | **0.03<sup>#</sup>** | 35.8 ± 9.4 | 36.4 ± 11.7 | 0.8<sup>#</sup> |
| **Sex (N)** | Male<br>(164) | Male<br>(257) | 0.7<sup>€</sup> | Male (61) | Male (46) | 0.3<sup>€</sup> |
| **Presence of Psychotic**<br>**symptoms (%)** | 86.3 | Not<br>applicable | Not<br>applicable | 86 | 89 | 0.5<sup>€</sup> |
| **Age at Onset (years)**<br>**(Mean ± SD)** | 21.5 ± 7 | Not<br>applicable | Not<br>applicable | 20.6 ± 6 | 22.4 ± 7.1 | **0.03<sup>#</sup>** |
| **Total number of**<br>**episodes (Mean ± SD)** | 6.3 ± 5 | Not<br>applicable | Not<br>applicable | 6.1 ± 3.6 | 8.1 ± 6.5 | 0.2<sup>#</sup> |

**Table 2:** Genotype and allele frequencies of *BDNF* (rs6265) in controls, cases and BD lithium response groups.

| Groups |  | Genotype |  |  | p-value | Allele |  | p-value |
| --- | --- | --- | --- | --- | --- | --- | --- | --- |
|  |  | GG | AG | AA |  | G<br>(Val) | A<br>(Met) |  |
| **Controls (N=484)** |  | 0.67<br>(323) | 0.29<br>(142) | 0.04<br>(19) | **0.02<sup>€</sup>** | 0.81<br>(788) | 0.19<br>(180) | **0.008<sup>€</sup>** |
| **Cases (N=301)** |  | 0.76<br>(229) | 0.21<br>(63) | 0.03<br>(9) |  | 0.87<br>(521) | 0.13<br>(81) |  |
| **Cases**<br>**(Lithium**<br>**response)** | **Responders (N=115)** | 0.61<br>(70) | 0.34<br>(39) | 0.05<br>(6) | **0.1<sup>¥</sup>** | 0.78<br>(179) | 0.22<br>(51) | **0.09<sup>€</sup>** |
|  | **Non-Responders**<br>**(N=75)** | 0.69<br>(52) | 0.31<br>(23) | 0.0<br>(0) |  | 0.85<br>(127) | 0.15<br>(23) |  |

€ Chi-square test;
¥ Fisher exact test.

## Discussion

We detected a significant difference in genotype frequency of Val66Met SNP between BD, and a matched healthy control sample. The frequency of the Val(G) allele was significantly higher in BD patients. As indicated earlier, there are population differences in the association of *BDNF* Val66Met polymorphism with BD risk. Our finding is consistent with the European population (De Luca et al., 2008; Frazier et al., 2014; Li et al., 2016), which indicate a positive association of the Val allele with BD risk. In the current study, the genotype and allele frequency of control subjects are similar to other reports from our group (Taj M J et al., 2017; Timothy, 2017, unpublished data (doctoral thesis)).

The frequency of the Val66Met polymorphism varies considerably, with both ethnicity and geography in the control population (Shen et al., 2018). It is interesting to note that the East Asian populations which showed association of the Met allele with BD have higher minor allele frequencies of 39-46%. On the other hand, Europeans with lower frequencies of 13-29% for the Met allele seem to show association with the Val allele. Further, our population which showed association with the Val allele also has a population frequency of 19% for the Met allele.

The Val allele of the *BDNF* Val66Met polymorphism has been found to be associated with reduced serum BDNF concentrations (Lang et al., 2009). The levels of BDNF in the circulating blood are reported to be lower in BD patients in comparison to healthy controls (Fernandes et al., 2015; Lin et al., 2016; Munkholm et al., 2016). BDNF is known to play an important role in promoting growth, development and survival of neuronal populations. It serves as a neurotransmitter modulator, and is involved in neuronal plasticity which is crucial for learning and memory (Bathina and Das, 2015). The neurotrophic hypothesis postulates that BD is associated with a lower activity of BDNF, and that lithium might exert protective actions by elevating BDNF (Sigitova et al., 2017).

We did not observe a significant association of the *BDNF* Val66Met polymorphism with response to lithium. As the sample size was underpowered to examine a pharmacogenetic association, we performed additional analysis of the extreme lithium response groups (total score ≥7 as lithium responder and score ≤3 as non-responder). There was no significant difference noted for this analysis. Our findings are consistent with some reports (Masui et al., 2006; Michelon et al., 2006; Wang et al., 2014), but in contrast to others which report a significant association of the Val allele with poor response to lithium (Dmitrzak-Weglarz et al., 2008; Rybakowski et al., 2007, 2012). Overall, the results of pharmacogenetic association studies of lithium with the *BDNF* Val66Met polymorphism have been inconsistent (Pagani et al., 2019; Pisanu et al., 2020).

We have attempted to address some of the anticipated limitations of the study. Older controls were chosen, as age matched controls could develop symptoms of the illness later in life. While the lithium treatment response was assessed retrospectively, we used standard methods as recommended by the Consortium on Lithium Genetics (ConLiGen) (Hou et al., 2016; Schulze et al., 2010). Since the sample was underpowered for pharmacogenetics, we additionally compared the extreme response groups.

Overall, our study suggests that the Val(G) allele of the *BDNF* Val66Met gene polymorphism is over-represented in BD in the Indian population. We did not, however, detect any association with lithium response. Genome-wide association studies across populations, to identify universal genetic mechanisms for BD, as well as underlying mechanisms for variations in response to lithium maybe the way forward.

## Data Availability

Data not available currently

## Acknowledgments

The authors would like to acknowledge Mr. Muralidharan Jayaraman, Lab technician for DNA isolation and Miss Varalakshmi, Lab assistant for technical assistance. We would like to thank the clinicians and staff at the NIMHANS; as well as the participants and their families for their co-operation.

